# Predicting low birth weight risks in pregnant women in Brazil using machine learning algorithms: Data from the Araraquara Cohort Study

**DOI:** 10.1101/2024.10.12.24315370

**Authors:** Audêncio Victor, Francielly Almeida, Sancho Pedro Xavier, Patrícia H.C. Rondó

## Abstract

**Background:** Low birth weight (LBW) is a critical factor linked to neonatal morbidity and mortality. Early prediction is essential for timely interventions. This study aimed to develop and evaluate predictive models for LBW using machine learning algorithms, including Random Forest, XGBoost, Catboost, and LightGBM.

**Methods:** Machine learning algorithms (Random Forest, XGBoost, Catboost, and LightGBM) were trained and evaluated using cross-validation and the SMOTE technique to correct class imbalance. Model performance was measured using the AUROC metric, and variable importance was analyzed with Shapley values to ensure model interpretability.

**Results:** The XGBoost model achieved the best performance with an AUROC of 0.94. Catboost and Random Forest also showed excellent results, confirming the effectiveness of these models in predicting LBW.

**Conclusion:** Machine learning, combined with SMOTE, proved to be an effective approach for predicting LBW. XGBoost stood out as the most accurate model, but Catboost and Random Forest also provided solid results. These models can be applied to identify high-risk pregnancies, improving perinatal outcomes through early interventions.

## Introduction

Birth weight is one of the primary determinants of newborn survival chances [1,2]. Low birth weight (LBW), defined as the birth of a baby weighing less than 2,500 grams, is a significant public health challenge associated with increased risk of infant mortality, neonatal morbidity, and chronic diseases in childhood and adulthood [3]. Several factors are considered in evaluating birth quality, including maternal nutritional status, prenatal care, and sociodemographic characteristics (BACCARAT DE GODOY MARTINS et al., 2016).

Recently, there has been an increase in LBW prevalence, predisposing children to respiratory diseases, growth retardation, heart conditions, and diabetes mellitus (MOREIRA; SOUSA; SARNO, 2018). LBW can result from preterm birth or inadequate fetal growth, often described as “small for gestational age” (SGA) to distinguish between immature infants and those with insufficient prenatal growth [6]. Intrauterine growth composition can influence the risk of cardiometabolic diseases in newborns, affecting both small and large for gestational age infants. The compensation of intrauterine growth through postnatal recovery or reduction may result in adverse outcomes (GÄTJENS et al., 2022). Various factors are identified as predictors of fetal growth, including intrauterine growth retardation, unfavorable socioeconomic conditions, inadequate prenatal care, low maternal education, maternal nutritional status, marital status, ethnicity/race, maternal weight, adolescent or advanced maternal age, urinary infections, and complications such as preeclampsia and bleeding during pregnancy (CHRISTINE et al., 2016; MOREIRA; SOUSA; SARNO, 2018).

In maternal and child health, artificial intelligence (AI), including machine learning (ML) algorithms, has been applied for outcome prediction and monitoring in perinatal health, offering new approaches for predictive modeling, diagnosis, early detection, and monitoring in perinatal health [8]. Machine learning is a subfield of AI aimed at extracting knowledge from large amounts of data, where algorithms are trained from previous examples [9]. This field has been on the rise in recent years due to the exponential increase in structured and unstructured data, also known as Big Data (BD), making ML approaches increasingly important in data analysis as traditional methods often rely on unrealistic assumptions [10,11].

In the context of maternal and child health, mobile health (mHealth) emerges as a promising AI application, particularly useful in prenatal care in low-resource settings [12]. The application of these algorithms can enhance the precision and reliability of predictions, contributing to early prevention and intervention strategies for fetal growth issues.

The goal of this study is to develop a predictive model for low birth weight (LBW) in pregnant women using machine learning algorithms. By applying these algorithms, we aim to improve the accuracy of predictions, thus enabling more effective strategies to address fetal growth problems early.

## Methods

This study is based on an empirical and quantitative application. Secondary data were analyzed from a longitudinal population-based cohort study conducted in Araraquara, São Paulo, Brazil, titled “Araraquara Cohort” [13]. The sample included women with a gestational age of ≤19 weeks who received prenatal care at Basic Health Units in Araraquara, São Paulo, Brazil. The pregnant women were followed quarterly throughout their pregnancy until the birth of their children between 2017 and 2022. Women with twin pregnancies and those who experienced miscarriage were excluded. In cases of fetal and stillbirth, only data from the pregnancy were considered.

The outcome of low birth weight was analyzed based on the dichotomous classification of birth weight, defined as low birth weight: < 2500 g and normal weight: ≥ 2500 g. The predictor variables are illustrated in Table 1.

**Table 1.**
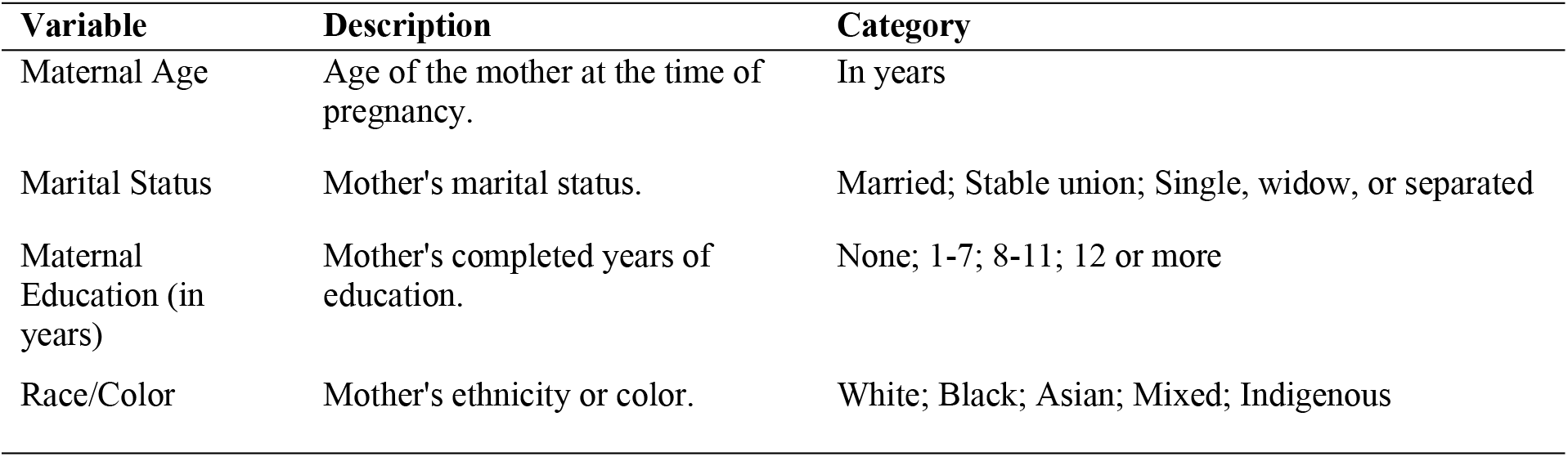
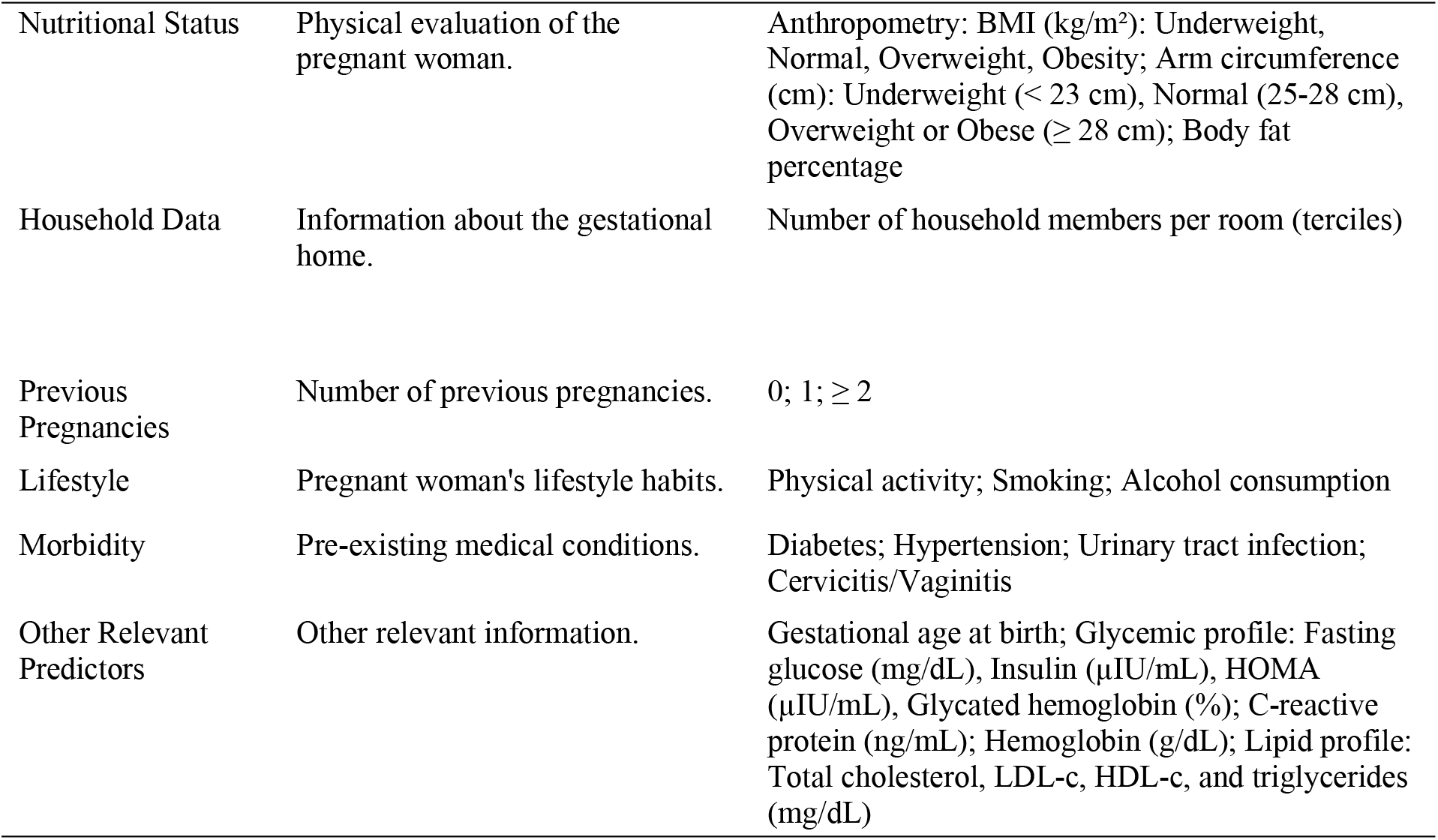
Description of model variables.

| Variable | Description | Category |
| --- | --- | --- |
| Maternal Age | Age of the mother at the time of pregnancy. | In years |
| Marital Status | Mother's marital status. | Married; Stable union; Single, widow, or separated |
| Maternal Education (in years) | Mother's completed years of education. | None; 1-7; 8-11; 12 or more |
| Race/Color | Mother's ethnicity or color. | White; Black; Asian; Mixed; Indigenous |
| Nutritional Status | Physical evaluation of the pregnant woman. | Anthropometry: BMI (kg/m<sup>2</sup>): Underweight, Normal, Overweight, Obesity; Arm circumference (cm): Underweight (< 23 cm), Normal (25-28 cm), Overweight or Obese (≥ 28 cm); Body fat percentage |
| Household Data | Information about the gestational home. | Number of household members per room (terciles) |
| Previous Pregnancies | Number of previous pregnancies. | 0; 1; ≥ 2 |
| Lifestyle | Pregnant woman's lifestyle habits. | Physical activity; Smoking; Alcohol consumption |
| Morbidity | Pre-existing medical conditions. | Diabetes; Hypertension; Urinary tract infection; Cervicitis/Vaginitis |
| Other Relevant Predictors | Other relevant information. | Gestational age at birth; Glycemic profile: Fasting glucose (mg/dL), Insulin (μIU/mL), HOMA (μIU/mL), Glycated hemoglobin (%); C-reactive protein (ng/mL); Hemoglobin (g/dL); Lipid profile: Total cholesterol, LDL-c, HDL-c, and triglycerides (mg/dL) |

The study was approved by the Research Ethics Committee with Human Subjects at the School of Public Health, University of São Paulo, prior to data collection, under protocol number CAEE: 59787216.2.0000.5421, opinion number 1.885.874. All participants signed the Informed Consent Form before participating. The participants were informed about the objectives of the study, the associated risks, and benefits, ensuring that their participation was entirely voluntary.

### Model Design

As shown in Figure 1, regarding the fetal growth outcome (LBW), different model specifications were tested to evaluate whether changes in strategy improved model performance. Models for the outcome were estimated independently, without information sharing between them. Before discussing the models, quantitative variables were normalized using the z-score separately for the training and test sets. All qualitative variables were treated through one-hot encoding, where each category was separately considered for this procedure. Additionally, pregnant women were excluded due to missing information, and variables with missing data below 20% were imputed by the mean.

**Figure 1.**
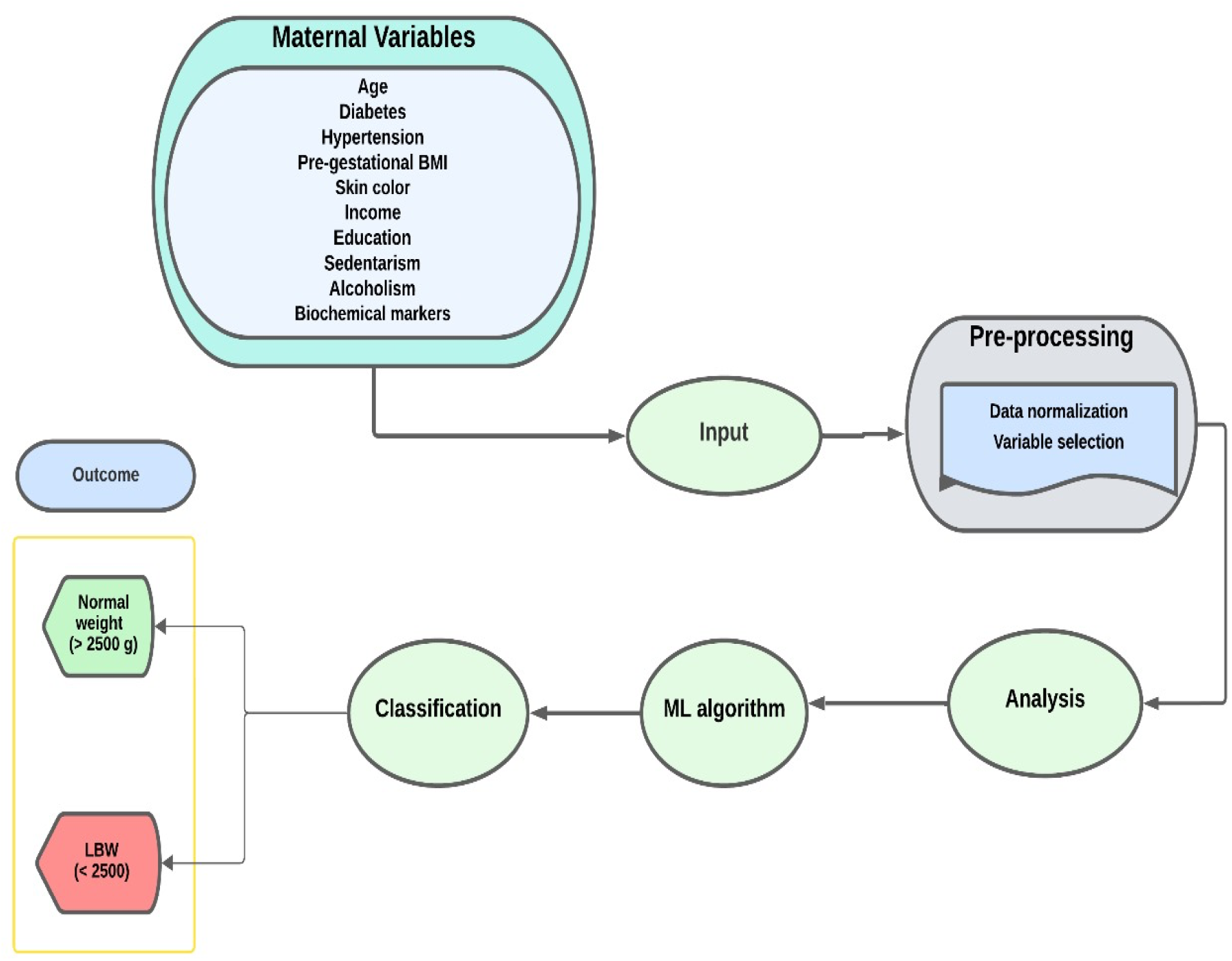
An example of the workflow diagram for classifying birth weight adequacy.

In this study, four different machine learning algorithms were tested: Catboost [14], Xgboost (CHEN e GUESTRIN, 2016), Lightgbm [16] and Random Forest. For Catboost, XGBoost, and LightGBM, the Python packages were used. For the remaining algorithms, the scikit-learn library was employed [17].Recent studies indicate that boosting algorithms represent the state-of-the-art for tabular data. They have shown high performance across a wide range of tasks, including classification (BORISOV et al., 2022; SHWARTZ-ZIV e ARMON, 2022).

The training set was conducted through 10-fold cross-validation using the Bayesian optimization strategy and RandomSearch to evaluate the performance of a machine learning model efficiently (BERGSTRA e YAMINS; COX, 2013). In cases of significant class imbalance, where the minority class represents less than 25% of the total outcomes, the Synthetic Minority Over-sampling Technique (SMOTE) was applied. Additionally, the Boruta method was employed for feature selection (KURSA e RUDNICKI, 2010). The best-performing models in the training set (80% of the data) were selected for evaluation in the test set (20%). The evaluation of ML algorithms was conducted in the test set using metrics such as area under the ROC curve (AUC-ROC), Matthews correlation coefficient (MCC), precision, recall, positive predictive value, negative predictive value, and F1-score. Additionally, the performance of the algorithms in the top 20% of high-risk patients (20% k-tops) was assessed using metrics such as true positive, false positive, precision, and recall. All analyses followed the Transparent Reporting of a multivariable prediction model for Individual Prognosis or Diagnosis (TRIPOD) guidelines [22]. The code developed for constructing the algorithms along with the original dataset, is available on Github https://github.com/Audency/Predictors-of-Low-Birth-Weight-using-Machine-Laerning-.git. This study was approved by the Research Ethics Committee of the School of Public Health, University of São Paulo (USP), prior to data collection, under CAEE number 59787216.2.0000.5421, opinion number 1.885.874. All participants provided informed consent, consistent with the principles outlined in the Helsinki Declaration.

## Results

### Maternal Characteristics

According to Table 2, the maternal characteristics of 1,579 pregnant women from the Araraquara cohort were evaluated. The women had an average age of 28.4 years, a height of 162 cm, a pre-pregnancy body mass index (BMI) of 24.7 kg/m^2^, and a gestational age of 39.3 weeks. Most women (88.4%) had an education level equal to or greater than 8 years, 53.7% were non-white, and 87.7% were married or in a stable relationship, with most having a family income of R$563 and being non-smokers. Only 5.0% of the women had diabetes, and 7.0% had hypertension. A total of 11.3% of the women had a urinary tract infection during pregnancy. CRP (C-reactive protein) levels were 3.3 mg/L (interquartile range: 1.4–7.8), and HOMA (homeostasis model assessment) values were 2.9 units (interquartile range: 1.3–6.1). Figure 2 shows the distribution of low birth weight in the Araraquara cohort, where 1,309 (91.2%) had normal birth weight, and 126 (8.8%) had low birth weight.

**Table 2.** Maternal characteristics of pregnant women in the Araraquara Cohort, Brazil (2021-2023)

| Variable | N (%) |
| --- | --- |
| **Age (years)** |  |
| < 20 | 154 (9.7) |
| 20 to 35 | 1,185 (74.9) |
| > 35 | 244 (15.4) |
| **Height (cm)** |  |
| < 153 | 534 (33.7) |
| 153-165 | 566 (35.8) |
| > 165 | 483 (30.5) |
| **Pre-pregnancy BMI (kg/m<sup>2</sup>)** |  |
| Underweight | 64 (4.0) |
| Normal weight | 1,084 (68.5) |
| Overweight | 450 (28.4) |
| Obesity | 48 (3.2) |
| **Arm circumference (cm)** |  |
| < 23 | 674 (42.6) |
| ≥ 23 | 919 (57.4) |
| **Gestational age (weeks)** | 39.3 (38.5–40.3) |
| **Maternal Education (years)** |  |
| ≤ 8 | 184 (11.6) |
| > 8 | 1,395 (88.4) |
| **Number of people per room** |  |
| ≤ 1 | 536 (33.9) |
| > 1 | 1,043 (66.1) |
| **Family income (R\$)** | |
| < 937 | 311 (32.5) |
| 937–1,866 | 563 (34.6) |
| > 1,866 | 666 (33.0) |
| **Race** |  |
| Non-white | 724 (46.3) |
| White | 833 (53.7) |
| **Marital Status** |  |
| Single | 135 (9.3) |
| Married or in a stable relationship | 1,359 (87.7) |
| Separated, divorced, or widowed | 198 (12.7) |
| **Smoking** |  |
| No | 1,434 (91.7) |
| Yes | 123 (7.9) |
| **Alcohol Consumption** |  |
| No | 1,235 (79.5) |
| Yes | 319 (20.5) |
| **Diabetes** |  |
| No | 1,479 (95.0) |
| Yes | 78 (5.0) |
| **Hypertension** |  |
| No | 1,445 (93.7) |
| Yes | 108 (7.0) |
| **Urinary Tract Infection** |  |
| No | 1,378 (88.7) |
| Yes | 175 (11.3) |
| **Candidiasis/Vaginitis** |  |
| No | 1,445 (91.9) |
| Yes | 108 (6.8) |
| **Number of previous births** |  |
| 0 | 764 (53.7) |
| ≥ 1 | 655 (43.9) |
| CRP (mg/L) | 3.3 (1.4–7.8) |
| HOMA (units) | 2.9 (1.3–6.1) |
| Glycated Hemoglobin (%) | 5.1 (4.7–5.6) |
| Fasting Glucose (mg/dL) | 74.3 (67.3–81.6) |
| Cholesterol (mg/dL) | 195 (168–224) |
| LDL (mg/dL) | 104 (86–123) |
| HDL (mg/dL) | 57 (47–68) |
| Triglycerides (mg/dL) | 94 (65–143) |

Note: Data are presented as mean and interquartile range (25th percentile - 75th percentile) or number (percentage). Abbreviations: BMI, Body Mass Index; LDL, Low-Density Lipoprotein; HDL, High-Density Lipoprotein; CRP, C-reactive Protein; HOMA, Homeostasis Model Assessment of Insulin Resistance.

**Figure 2.**
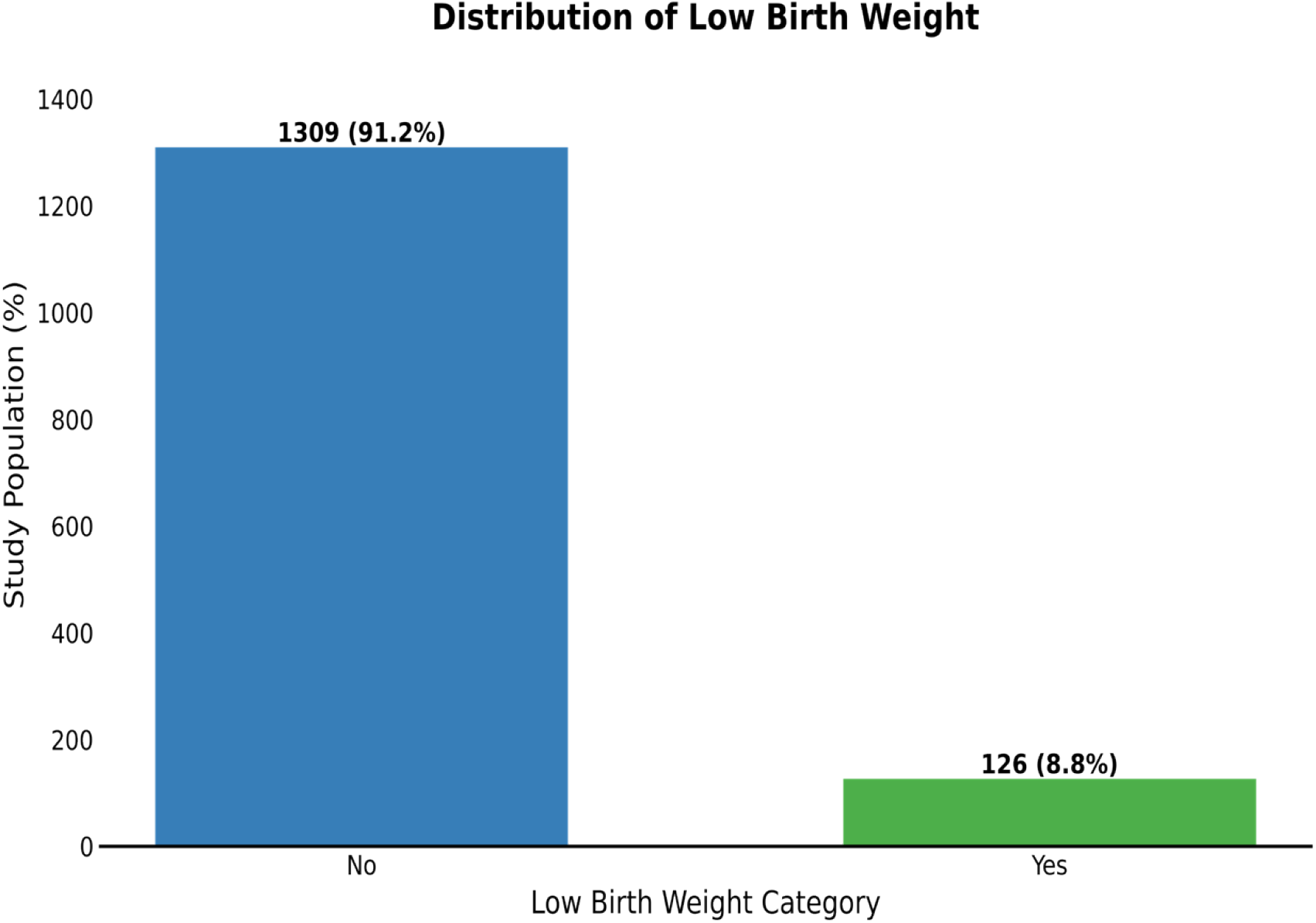
Distribution of LBW

## Results

### Performance of Machine Learning Models

In this study, we evaluated the predictive capacity of various machine learning models, including Random Forest, XGBoost, LightGBM, and Catboost, to predict LBW in neonates. The analysis focused on multiple performance metrics such as AUROC, accuracy, precision, recall, F1-score, and the MCC to provide a comprehensive evaluation of the ability of these models to classify cases of low birth weight.

The results of the machine learning models’ evaluation for LBW prediction are summarized in Table 3 and Figure 3. The XGBoost model showed the best performance with an AUROC of 0.941, demonstrating excellent discrimination between neonates with normal and low birth weight. Catboost followed closely with an AUROC of 0.939, while Random Forest and LightGBM achieved AUROCs of 0.938 and 0.937, respectively.

**Table 3.**
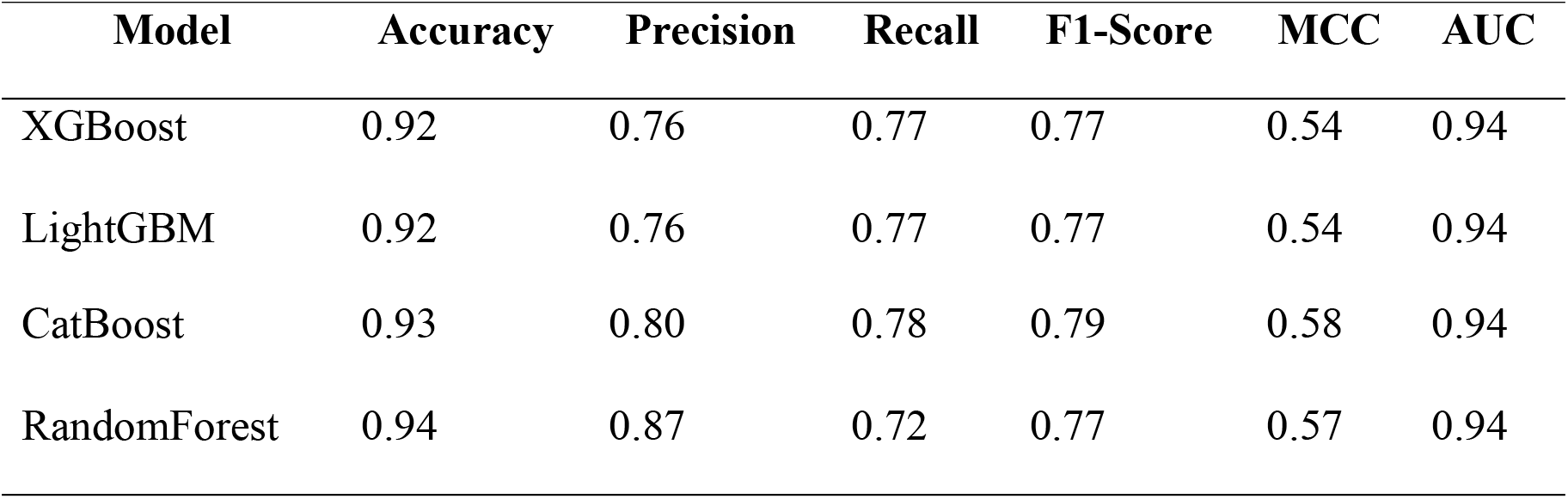
Test Set Performance comparison for LBW Weight Classification Models.

**Figure 3.**
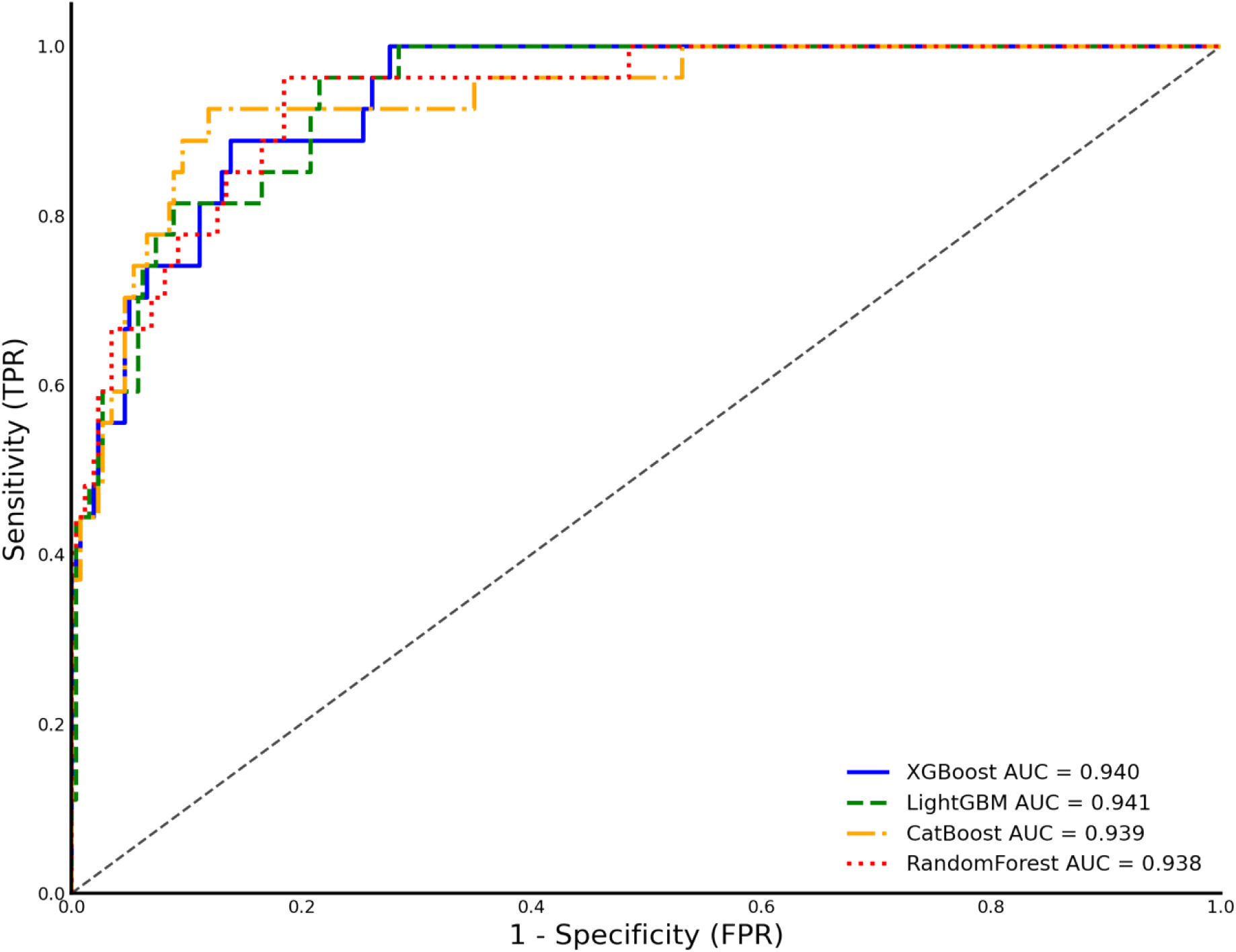
Performance by ROC Curve of ML (Random Forest, XGBoost, LightGBM, and Catboost) for LBW Prediction.

In addition to AUROC, other performance metrics were considered. Random Forest exhibited the highest overall accuracy (0.94), while Catboost provided the best balance between precision (0.80) and recall (0.78), resulting in an F1-score of 0.79 (Table 3). The ROC curves in Figure 3 visually illustrate the similar performance between the models, with all curves approaching the top-left corner, indicating high discriminatory capacity.

### Variable importance for LBW Prediction

Figure 4 shows the importance of the predictors for LBW using Shapley values for the best-performing model, XGBoost. The most important variable identified was gestational weight gain, standing out as the factor with the most influence on predicting low birth weight. Following this, maternal marital status and the absence of regular physical activity during pregnancy were significant predictors. Other factors contributing substantially included maternal race, parity, and fewer prenatal visits, underscoring the importance of socioeconomic and behavioral variables. Variables such as smoking and alcohol consumption during pregnancy, although important predictors, appeared with less relevance compared to the previously mentioned factors. S.1, illustrates the strength of variable contributions to the prediction of LBW using Shapley values in the XGBoost model. The most influential variables, shown in the graphs, provide a detailed view of how each factor contributes to increasing or reducing the risk of LBW.

**Figure 4.**
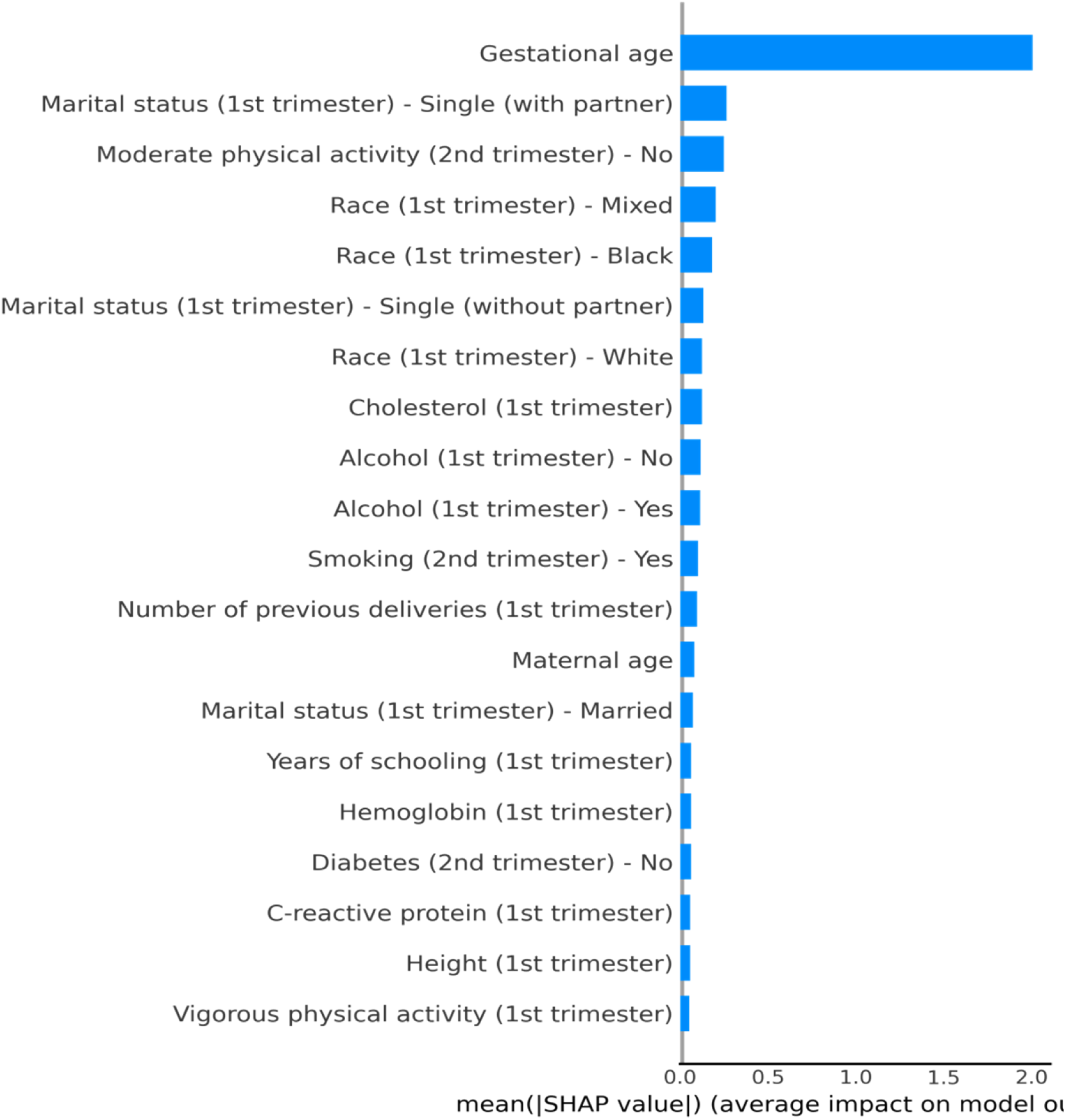
Predictors of LBW (Shapley Variable Importance) for the Best Model - XGBoost.

## Discussion

The findings from this study confirm and expand the evidence that machine learning (ML) models, such as Random Forest, XGBoost, LightGBM, and Catboost, are effective for predicting low birth weight (LBW) in neonates. Among the models tested, XGBoost exhibited the best performance with an AUROC of 0.941, which places it as the most effective algorithm. This outcome aligns with previous studies that have explored ML approaches in predicting LBW (NAIMI; PLATT e LARKIN, 2018a; SCHMIDT et al., 2022; WŁODARCZYK et al., 2021). The use of ML in LBW prediction is gaining traction, as these models demonstrate significant potential across various clinical and public health applications.

In agreement with the current literature, our study corroborates the efficacy of ML algorithms in maternal-fetal health. Previous studies have underscored the importance of selecting the most suitable ML algorithm, given the variations in performance and reliability among different models [26]. For example, a study published in BMC Pregnancy and Childbirth evaluated the performance of eight different ML algorithms for predicting LBW. It found that deep learning (AUROC: 0.86), random forest classification (AUROC: 0.79), and extreme gradient boost classification (AUROC: 0.79) performed best in distinguishing LBW from normal birth weight across a cohort of 8,853 births, where 1,280 resulted in LBW [27].

Our study further supports the finding that boosting algorithms such as XGBoost and Catboost are particularly powerful for tabular datasets, as seen in our cohort. These models outperform more traditional methods like logistic regression and are advantageous in dealing with complex non-linear relationships among predictors[18,19,28]. Studies like that of Pollob et al. (2022), which used ML to predict LBW in Bangladesh, similarly demonstrated that ensemble and boosting models provide more accurate predictions than classical statistical models, with LBW rates around 16.2% in their study population. Key risk factors included region, education, wealth index, and height, consistent with our findings on the importance of gestational weight gain, race, and socioeconomic status in predicting LBW.

The use of Shapley values in our study provided additional insights into the interpretability of the models, allowing for a more granular understanding of how each variable contributes to the prediction of LBW. This aspect is particularly important in clinical applications where transparency and explainability of the models are crucial for their adoption by healthcare professionals. For example, the Shapley analysis in our study indicated that gestational weight gain had the strongest influence on LBW predictions, followed by maternal race and prenatal care visits. These findings mirror global trends in LBW prediction, where maternal health, nutrition, and prenatal care are recognized as key determinants of birth outcomes.

Our findings also align with the work of Patterson et al. (2023), who developed a predictive model for LBW in low- and middle-income countries. Their study found that socioeconomic factors, including maternal education and access to prenatal care, significantly contributed to the risk of LBW, similar to our results from the Araraquara cohort. Both studies emphasize the importance of early identification of high-risk pregnancies through predictive models, especially in resource-limited settings where timely interventions can significantly improve neonatal outcomes.

The clinical implications of our findings are significant. By accurately identifying pregnancies at high risk for LBW, these ML models could enable healthcare providers to implement early interventions, such as nutritional supplementation, increased prenatal visits, or targeted counseling on lifestyle modifications. Such interventions could mitigate the risks associated with LBW, including neonatal mortality, morbidity, and long-term health consequences like developmental delays and chronic conditions. This is particularly relevant in low- and middle-income countries, where LBW rates are higher, and healthcare resources are often limited[29]

## Limitations and future directions

While the results of this study are promising, there are several limitations to consider. First, the cohort used in this study was from a specific region in Brazil, which may limit the generalizability of the findings to other populations. Future research should aim to validate these models across different regions and populations to ensure their broader applicability. Additionally, while our models demonstrated high predictive accuracy, further research is needed to assess their integration into clinical workflows and their potential impact on perinatal care. For example, future studies could explore the use of these models in combination with mobile health (mHealth) technologies to improve prenatal care in low-resource settings.

Moreover, although we employed robust techniques such as SMOTE to handle class imbalance and Shapley values to interpret model predictions, the models still require validation in real-world clinical environments. The practical deployment of ML models in healthcare settings involves challenges related to data privacy, model fairness, and bias mitigation, which should be thoroughly addressed before these models can be widely adopted.

## Conclusion

This study successfully developed and evaluated machine learning models, including XGBoost, Catboost, LightGBM, and Random Forest, for predicting low birth weight in neonates. The XGBoost model demonstrated the highest predictive performance, with excellent discrimination between neonates at risk of LBW and those with normal birth weight. The application of these models in clinical practice has the potential to improve early detection of high-risk pregnancies, enabling timely and personalized interventions that could significantly improve neonatal outcomes.

Given the increasing global focus on maternal and neonatal health, these findings hold important implications for both clinical practice and public health policy. The integration of machine learning models into prenatal care systems could offer a transformative approach to preventing adverse birth outcomes, particularly in low-resource settings where LBW remains a critical challenge.

## Data Availability

Data Availability Statement Response The data from the Araraquara cohort cannot be shared publicly due to ethical and confidentiality concerns. The cohort includes sensitive personal health information that is protected by the Brazilian General Data Protection Law (Lei Geral de Proteção de Dados – LGPD). Additionally, the participants provided informed consent for their data to be used only in the context of specific research projects and not for public distribution. However, data access can be requested through the Research Ethics Committee of the School of Public Health, University of São Paulo (USP), upon approval. Researchers interested in accessing the data should contact the committee via, provided they meet the necessary criteria for handling confidential and sensitive data.

## Acknowledgments

The authors gratefully acknowledge the professionals, undergraduate, and graduate students who collaborated in the data collection for the Araraquara cohort. We also thank the editors and reviewers for their valuable suggestions and comments to improve our manuscript. Additionally, we acknowledge the São Paulo Research Foundation (FAPESP) for financial support (grant number 2015/03333-6) and the principal author’s scholarship (grant number 2023/07936-3).

## Author Contributions

**Conceptualization:** Audê ncio Victor and Francielly Almeida.

**Data curation:** Audê ncio Victor.

**Formal analysis:** Audê ncio Victor.

**Investigation:** Audê ncio Victor, Sancho Pedro Xavier.

**Methodology:** Audê ncio Victor, Francielly Almeida, Patrícia H.C. Rondó.

**Supervision:** Francielly Almeida and Patrícia H.C. Rondó

**Writing – original draft:** Audê ncio Victor and Sancho Pedro Xavier.

**Writing – review & editing:** Audê ncio Victor, MPH, Francielly Almeida, Sancho Pedro Xavier, Patrícia H.C. Rondó.

